# Depressive disorders are associated with increased peripheral blood cell deformability: A cross-sectional case-control study (Mood-Morph)

**DOI:** 10.1101/2021.07.01.21259846

**Authors:** Andreas Walther, Anne Mackens-Kiani, Julian Eder, Maik Herbig, Christoph Herold, Clemens Kirschbaum, Jochen Guck, Lucas Daniel Wittwer, Katja Beesdo-Baum, Martin Kräter

## Abstract

**Background:** Pathophysiological landmarks of depressive disorders are chronic low-grade inflammation and elevated glucocorticoid output. Both can potentially interfere with cell membrane bending and cell function, suggesting altered cell morpho-rheological properties like cell deformability and other cell mechanical features in depressive disorders.

**Method:** We performed a cross-sectional case-control study using image-based morpho-rheological characterization of unmanipulated blood samples facilitating real-time deformability cytometry (RT-DC). Sixty-nine pre-screened individuals at high-risk for depressive disorders and 70 matched healthy controls were included and clinically evaluated by Composite International Diagnostic Interview. Facilitating deep learning on blood cell images, major blood cell types were classified and morpho-rheological parameters such as cell size and cell deformability of every individual cell was quantified.

**Results:** We found peripheral blood cells to be more deformable in patients with depressive disorders compared to controls, while cell size was not affected. Lifetime persistent depressive disorder was associated with an increased cell deformability in monocytes and neutrophils, while in current persistent depressive disorder erythrocytes deformed more. Lymphocytes were more deformable in current major depressive disorder, while for lifetime major depressive disorder no differences could be identified.

**Conclusions:** This is the first study analyzing morpho-rheological properties of entire blood cells and highlighting depressive disorders and in particular persistent depressive disorders to be associated with increased blood cell deformability. While all major blood cells tend to be more deformable, lymphocytes, monocytes, and neutrophils are mostly affected. This indicates that immune cell mechanical changes occur in depressive disorders, which might be predictive for persistent immune response.

## Introduction

Depressive disorders including major depressive disorder (MDD) and persistent depressive disorder (PDD; formerly dysthymia) are the leading causes of disability worldwide (World Health Organisation 2017). To diagnose MDD a two-week phase is required. During this phase at least one of the two cardinal symptoms “depressive mood” or “anhedonia” in combination with four or more of seven other symptoms (e.g. changes in appetite, insomnia/hypersomnia, increased fatigue, feelings of worthlessness) have to be present for most of the day and must cause functional impairment. PDD is diagnosed based on a period of depressive mood over two-years in combination with at least two of six additional symptoms similar as for MDD ^2^. To date, physiological manifestations only play a theoretical role for diagnostics. This is due to the fact that the pathophysiology of depressive disorders remains insufficiently understood. The two most consistent and salient physiological abnormalities are a hyperactive hypothalamus-pituitary-adrenal (HPA) axis and chronic low-grade inflammation associated with elevated cortisol and proinflammatory cytokine levels, respectively ^3^. In line with this, an increased lymphocyte count has been identified in MDD and PDD ^4^. Additionally, increased neutrophil and monocyte counts were described, suggesting distinct subgroups of patients based on blood cell immunophenotyping and pro-inflammatory cytokine and protein levels ^5^. Blood cells represent the first target of increased cortisol levels and chronic low-grade inflammation, crucially affecting lipid metabolism underlying cell membrane formation. Altered cell lipid composition is leading to increased membrane bending and destabilization ^6–9^. Therefore, we hypothesize depressive disorders to be associated with altered peripheral blood cell function, which might be represented by the cells morpho-rheological properties ^6^.

Blood is a poly-disperse suspension of a number of different cell types, representing multiple functions from metabolite transport to overall blood flow. The morpho-rheological properties including cell mechanical features or cell size of each cell can be predictive of its specific physiological or pathological function ^10^. It was recently highlighted, that the assessment of the blood cell mechanical status, measured by cell deformability under constant shear stress, is appropriate to detect and classify human disease conditions ^10^. Furthermore, proof-of-concept studies have used optical traps ^11^, atomic force microscopy ^12,13^, or micropipette aspiration ^14^ to show immune cell mechanical alterations during physiological and pathological conditions. Most likely due to the overall predominance of erythrocytes, a correlation of blood cell mechanics and mental disorders has so far only been examined by measuring erythrocyte deformability ^15,16^. However, immune cells seem to be more likely effectors of increased cortisol levels and increased chronic low-grade inflammation. Thus, a progress towards clinical application has yet not been achieved, potentially due to the lack of measurement throughput with only a couple of hundred cells per hour ^17^.

Here, we used state of the art real-time deformability cytometry (RT-DC) together with artificial intelligent-based image processing in order to overcome the throughput limitations (Figure 1). We measured the morpho-rheological properties of more than 16 × 10^6^ single blood cells of 69 individuals at high-risk for depressive disorders and 70 matched healthy controls (HCs). RT-DC facilitates microfluidics and high-throughput imaging to assess up to 1,000 cells per second. Based on the cell image, RT-DC quantifies multiple parameters including the cell’s deformability under shear stress and cell size without the need for blood preparation, like cell staining or erythrocyte depletion ^18^. The observed deformability is dependent on the cell’s mechanical properties. These properties are representative for the molecular composition and the cytoskeletal state ^19,20^, which are arguably the most crucial aspects to fulfill tissue and cell-specific functionality. Thus, we argue that the precise control of mechanical features of blood cells is indispensable to keep physical and psychological homeostasis. Thus, cell mechanical properties potentially comprise crucial pathophysiological information in mental disorders and particular in depressive disorders.

**Figure 1:**
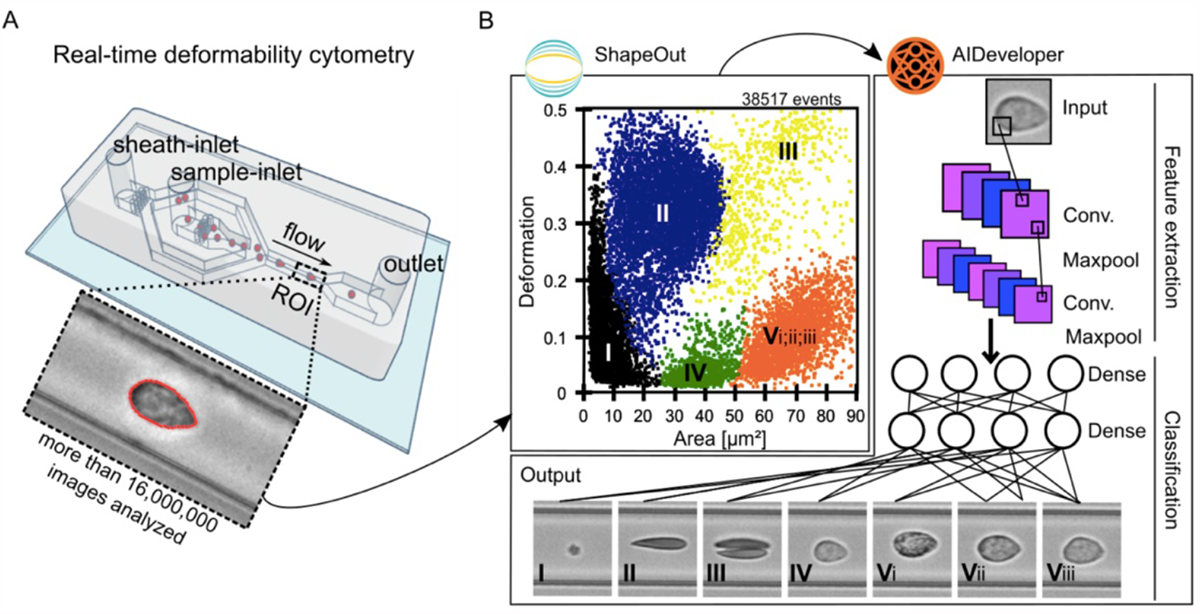
Real-time deformability cytometry and subsequent AI-based classification of blood cells. (A) A schematic illustration of an RT-DC measurement chip is shown. Whole blood was resuspended in measurement buffer (CellCarrierB), drawn in a syringe and connected to the sample-inlet. CellCarrierB was used as a sheath fluid within a second syringe and sample and sheath were flushed through the chip at a ratio of flow rates of 1:3 under constant flow (0.06 µL/s). The chip was mounted to an inverted microscope and an image of every cell was recorded at the end of a 600 µm long constriction cannel. (B) Using ShapeOut, an open source software-tool, data was plotted. The dot-plot shows a measurement of 38,517 blood cells plotted in cell size (projected area [µm^2^]) and cell deformability. For better representation, the ratio of leucocytes (IV, V_i_-_iii_) to erythrocytes and thrombocytes (I-III) is artificially increased. In order to identify the different blood cell types, the images where imported to AIDeveloper an open source software-tool to train, evaluate and apply neural networks for image classification. A neural net based on the LeNet5 architecture, readily trained for the classification of blood cells, was loaded into AID and used to classify I = thrombocytes, II = erythrocytes, III = erythrocyte doublets, IV = lymphocytes, V_i_ = eosinophils, V_ii_ = neutrophils and V_iii_ = monocytes.^24^ Finally, the mean values for cell deformability and cell size were extracted for every cell type individually.

## Method

### Study design and setting

The study entitled Mood-related morpho-rheological changes in peripheral blood cells (Mood-Morph) included a pre-screening to select participants suffering from depressive disorders and healthy control subjects matched by age and sex, followed by a cross-sectional case-control study. Study visits included a clinical diagnostic interview, psychometric testing, and blood sampling.

### Participants

Recruitment was performed from the participant pool of the large prospective cohort study on stress-related mental disorders (Dresden Burnout Study [DBS]) with respect to depression scores (measured with the Patient Health Questionnaire [PHQ-9]) in the most recent examination wave from October to December 2018. Subjects with a score > 10 (high risk of depression) as well as age- and sex-matched subjects with a score < 5 (low risk of depression) were invited. This procedure’s aim was to achieve a sample of participants suffering from depressive disorders and a matched sample of healthy control subjects (Figure 2).

**Figure 2:**
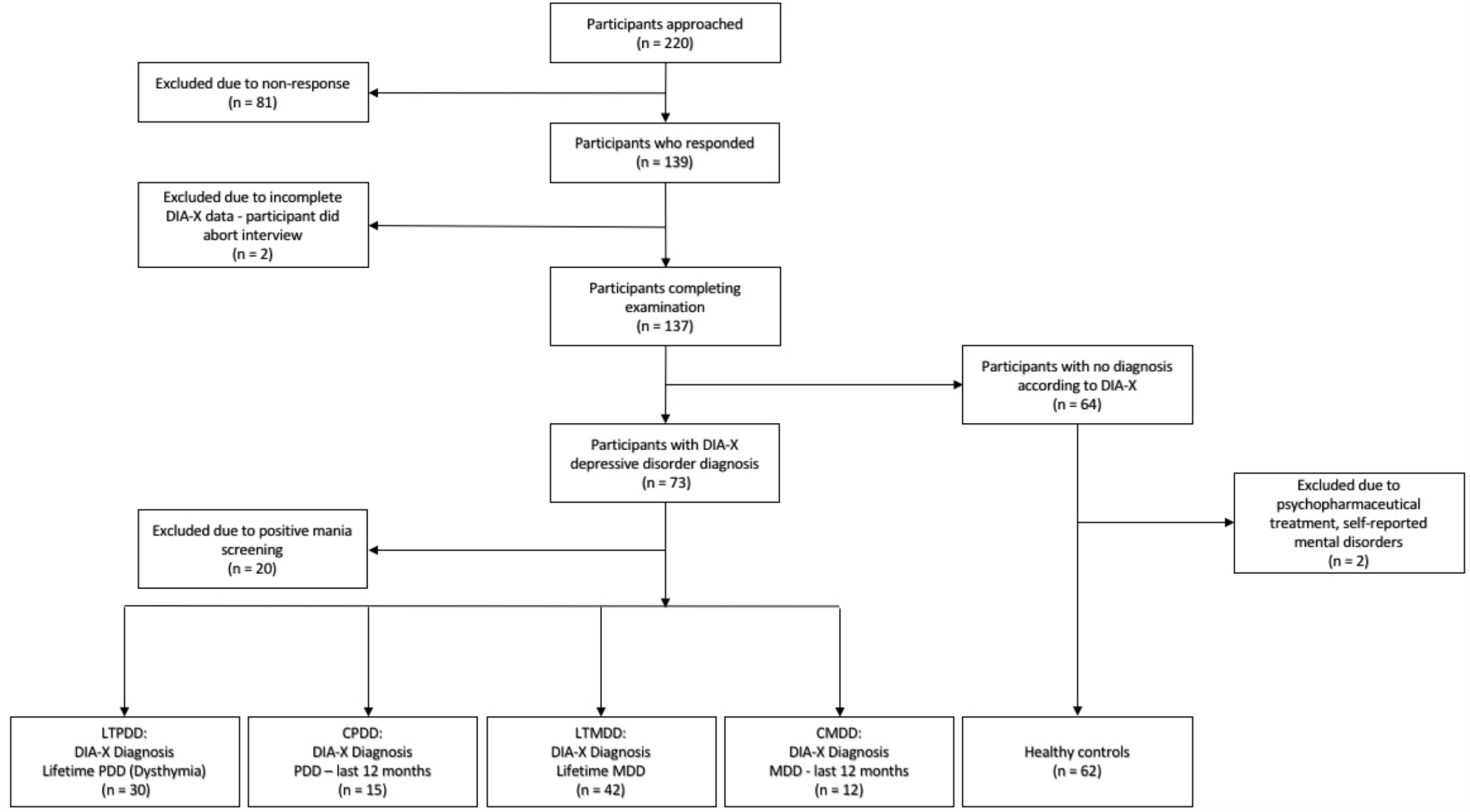
Sample flow leading to relevant case-control groups. DIA-X-5: The DIA-X-5/Composite International Diagnostic Interview (DIA-X-5/CIDI) is a standardised clinical interview for the assessment of mental disorders, LTPDD: Lifetime persistent depressive disorder, CPDD: Current persistent depressive disorder, LMDD: Lifetime major depressive disorder, CMDD: Current major depressive disorder.

### Procedures

Depressive symptom severity as well as general health were measured online via the DBS-homepage one week prior to examination using the depression section of the PHQ-9 and the Short Form Health Questionnaire (SF-12) ^21,22^. The PHQ is a self-report questionnaire for the assessment of common mental disorders with the PHQ-9 being the module designed for the measurement of depression severity. The SF-12 measures physical and psychological health-related quality of life.

During examination, participants completed a subject questionnaire regarding age, sex, weight, height, medication, drug use, psychopharmacological treatment and psychotherapeutic treatment. After completion of the questionnaire the depression section of the Composite International Diagnostic Interview (DIA-X-5/CIDI) was conducted by trained interviewers ^23^. The DIA-X-5 is a standardized clinical interview for the assessment of mental disorders. A mania-screening questionnaire, consisting of the initial questions of the DIA-X-5 mania section, was conducted to detect any lifetime (hypo-) mania-symptoms. Lifetime and current diagnoses were subsequently generated for PDD and MDD according to DSM-5 criteria ^2^. Diagnosis of current PDD or MDD was based on past twelve month assessment. Subsequently, a 20 µl capillary blood sample was extracted from the fingertip using a safety lancet. The blood was immediately diluted with 380 µl of measurement buffer (CellCarrierB, Zellmechanik Dresden, Germany) in a microcentrifuge tube.

All subjects signed consent forms regarding data privacy, clinical data collection and saving, and blood extraction. Subjects received 15 € compensation for expenses. Duration of the examination was approximately one hour. The study was approved by the local ethics committee of the Dresden University of Technology.

After examination, blood-samples were transferred to the Department of Cellular Machines at the Biotechnology Center of the TU Dresden, where the samples were measured using an RT-DC device.

### Real-time deformability cytometry (RT-DC)

A 20 µl blood drop was taken from study participants by finger pricking using a lancet (Safety-Lancet Normal 21, Sarstedt, Nümbrecht, Germany) and harvested in a capillary (Minivette POCT, 20 µl, Sarstedt, Nümbrecht, Germany). Blood was immediately resuspended in 380 µl RT-DC measurement buffer containing 0.6% methylcellulose (CellCarrierB; Zellmechanik Dresden, Germany), maintained at room temperature, and measured within three hours according to a protocol published elsewhere ^10^. In brief, blood was flushed through a microfluidic channel constriction 20 µm⨯ 20 µm in cross section (Flic20, Zellmechanik Dresden, Germany) by applying a constant flow rate. An image of every measured blood cell was taken by a high-speed camera (Figure 1A) and beside other parameters cell deformability and projected area (cell size) were calculated ^18^. RT-DC measurements were controlled by the acquisition software Shape-In2 (Zellmechanik Dresden, Germany). The different blood cell types were classified by utilizing artificial intelligence-based image classification as published elsewhere ^24^ and mean values for cell deformability and cell size of every donor and blood cell type were extracted (Figure 1B).

### Statistical analysis

Data was analyzed using R 3.4.3 (R Core Team, 2017). Two-tailed independent *t*-tests and Mann-Whitney *U* tests were performed to compare deformability and cell size (projected area [µm^2^]) of each blood cell type between healthy controls and depressed individuals. Data was checked for normality using Shapiro-Wilk-tests. Moderating confounders (sex, age, BMI, psychopharmaceutical intake, medication category) were either adjusted for or tested using two-tailed independent t-tests and univariate ANOVA.

### Role of funding source

The funding source had no influence on the design of the study, subject recruitment, collection of the data, statistical analysis, interpretation of the data, or the writing of the manuscript.

## Results

### Case-control distribution for the group contrasts

A total of 139 pre-screened individuals scoring above 10 (*n* = 69) or below 5 (*n* = 70) in the PHQ-9 were examined in the study. Individuals meeting a positive screen for mania were excluded from PDD and MDD groups. Individuals reporting psychopharmaceutical treatment or mental disorders in the subject questionnaire were excluded from the healthy control group. Exclusion criteria and the DIA-X-5 diagnosis led to the following groups: Lifetime persistent depressive disorder (LTPDD) (*n* = 30), current persistent depressive disorder (CPDD) (*n* = 15), lifetime major depressive disorder (LTMDD) (*n* = 42), current major depressive disorder (CMDD) (*n* = 12) and healthy controls (HC) (*n* = 62). PDD and MDD groups partially overlap. Each group of participants suffering from depression was compared to an age- and sex-matched healthy control group. For detailed sample characteristics see Table 1.

**Table 1.**
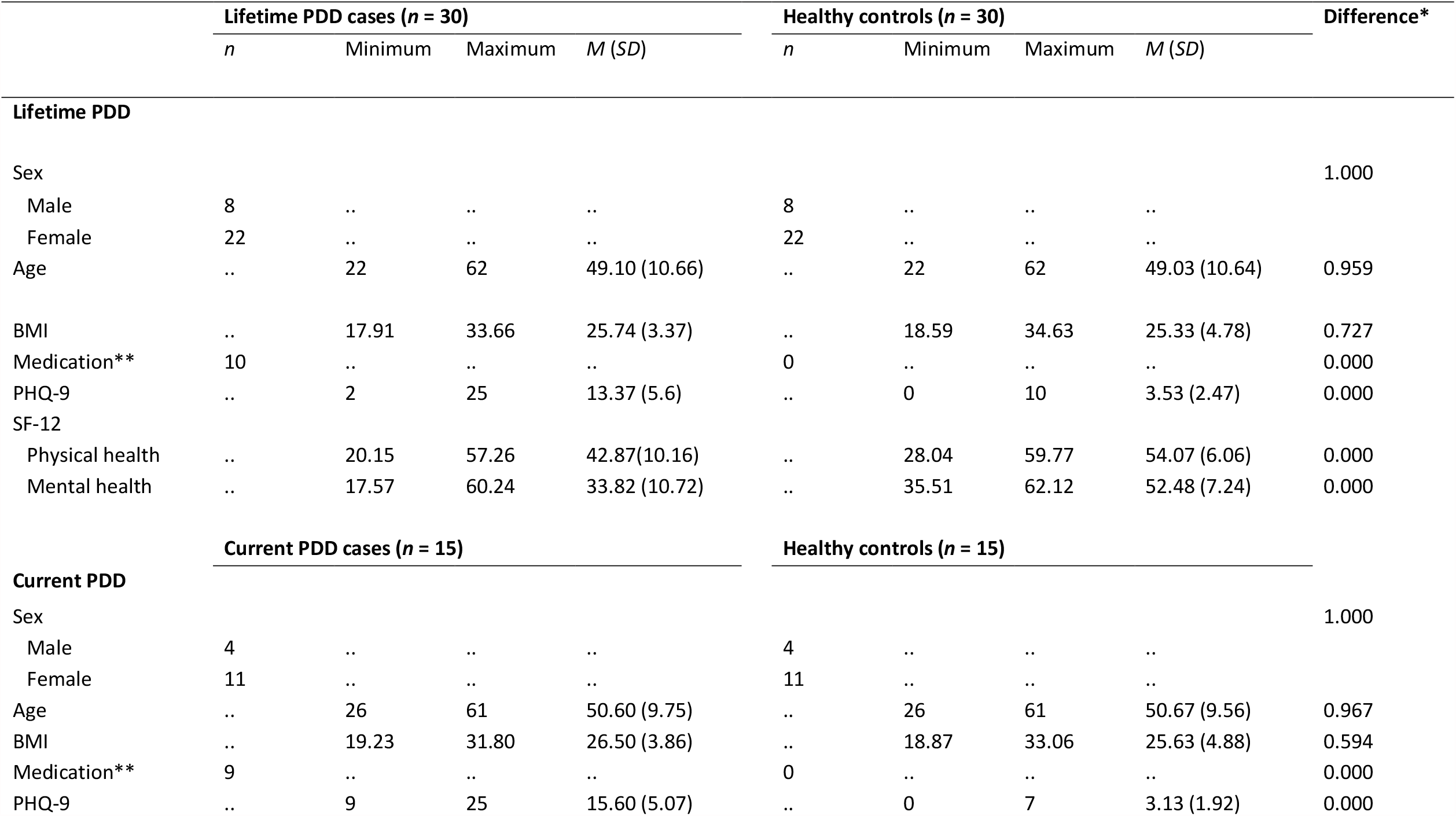
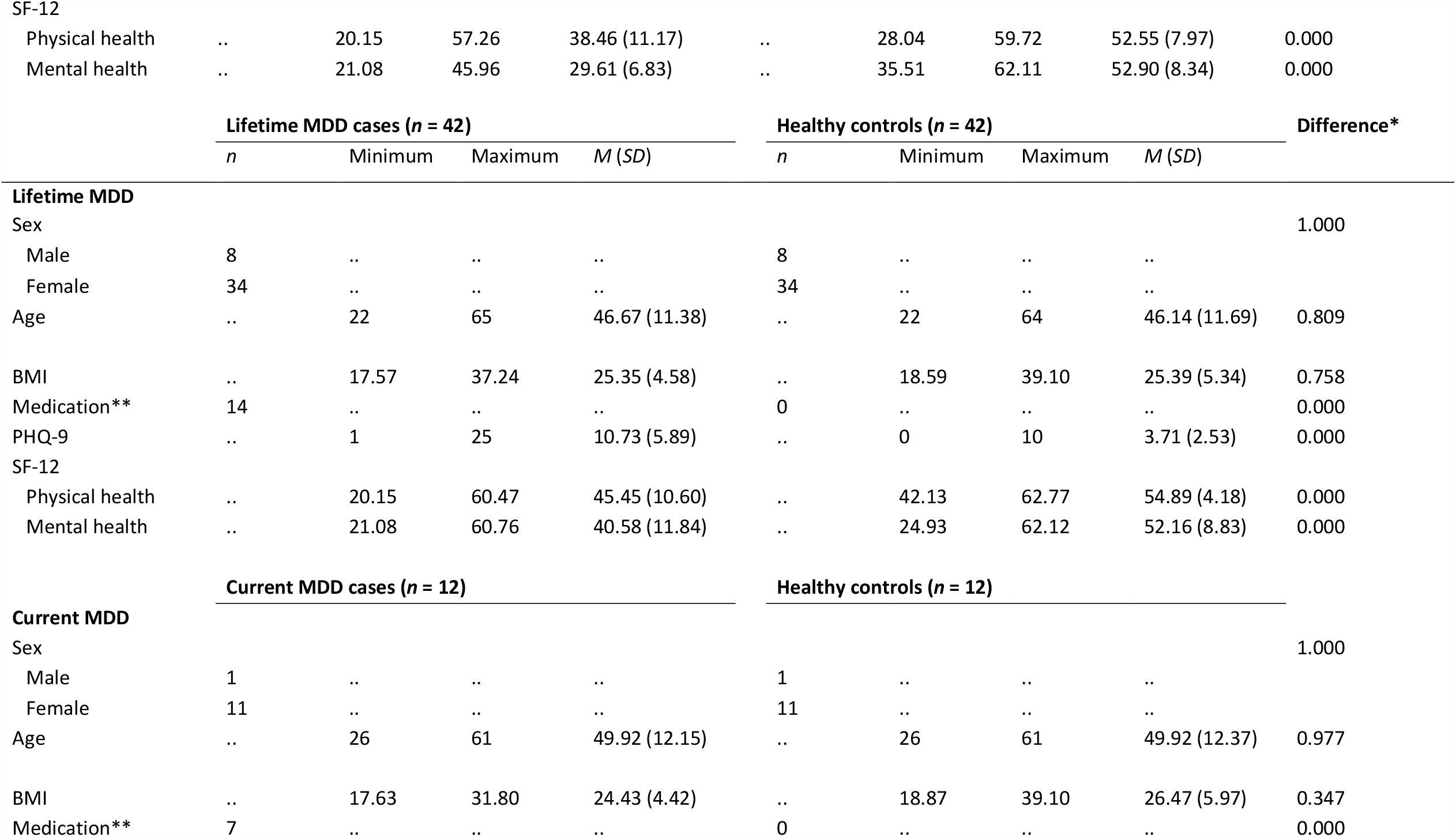
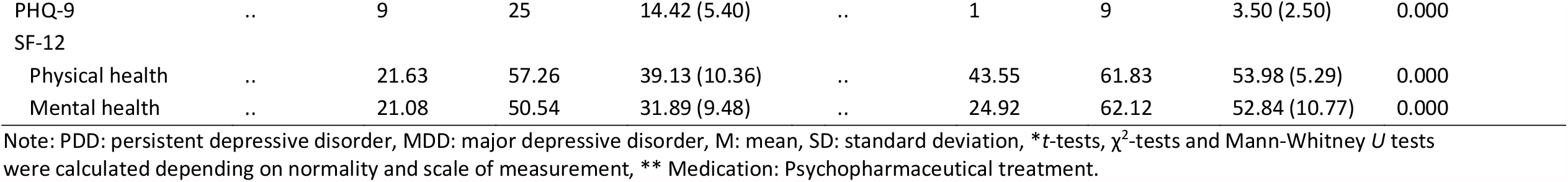
Sample characteristics according to different diagnostic groups.

|  | Lifetime PDD cases (*n* = 30) |  |  |  | Healthy controls (*n* = 30) |  |  |  | Difference* |
| --- | --- | --- | --- | --- | --- | --- | --- | --- | --- |
|  | *n* | Minimum | Maximum | *M* (*SD*) | *n* | Minimum | Maximum | *M* (*SD*) |  |
| Lifetime PDD |  |  |  |  |  |  |  |  |  |
| Sex |  |  |  |  |  |  |  |  | 1.000 |
| Male | 8 | .. | .. | .. | 8 | .. | .. | .. |  |
| Female | 22 | .. | .. | .. | 22 | .. | .. | .. |  |
| Age | .. | 22 | 62 | 49.10 (10.66) | .. | 22 | 62 | 49.03 (10.64) | 0.959 |
| BMI | .. | 17.91 | 33.66 | 25.74 (3.37) | .. | 18.59 | 34.63 | 25.33 (4.78) | 0.727 |
| Medication** | 10 | .. | .. | .. | 0 | .. | .. | .. | 0.000 |
| PHQ-9 | .. | 2 | 25 | 13.37 (5.6) | .. | 0 | 10 | 3.53 (2.47) | 0.000 |
| SF-12 |  |  |  |  |  |  |  |  |  |
| Physical health | .. | 20.15 | 57.26 | 42.87(10.16) | .. | 28.04 | 59.77 | 54.07 (6.06) | 0.000 |
| Mental health | .. | 17.57 | 60.24 | 33.82 (10.72) | .. | 35.51 | 62.12 | 52.48 (7.24) | 0.000 |
|  | Current PDD cases (*n* = 15) |  |  |  | Healthy controls (*n* = 15) |  |  |  |  |
| Current PDD |  |  |  |  |  |  |  |  |  |
| Sex |  |  |  |  |  |  |  |  | 1.000 |
| Male | 4 | .. | .. | .. | 4 | .. | .. | .. |  |
| Female | 11 | .. | .. | .. | 11 | .. | .. | .. |  |
| Age | .. | 26 | 61 | 50.60 (9.75) | .. | 26 | 61 | 50.67 (9.56) | 0.967 |
| BMI | .. | 19.23 | 31.80 | 26.50 (3.86) | .. | 18.87 | 33.06 | 25.63 (4.88) | 0.594 |
| Medication** | 9 | .. | .. | .. | 0 | .. | .. | .. | 0.000 |
| PHQ-9 | .. | 9 | 25 | 15.60 (5.07) | .. | 0 | 7 | 3.13 (1.92) | 0.000 |
| SF-12 |  |  |  |  |  |  |  |  |  |
| Physical health | .. | 20.15 | 57.26 | 38.46 (11.17) | .. | 28.04 | 59.72 | 52.55 (7.97) | 0.000 |
| Mental health | .. | 21.08 | 45.96 | 29.61 (6.83) | .. | 35.51 | 62.11 | 52.90 (8.34) | 0.000 |

|  | Lifetime MDD cases (*n* = 42) |  |  |  | Healthy controls (*n* = 42) |  |  |  | Difference* |
| --- | --- | --- | --- | --- | --- | --- | --- | --- | --- |
|  | *n* | Minimum | Maximum | *M* (*SD*) | *n* | Minimum | Maximum | *M* (*SD*) |  |
| Lifetime MDD |  |  |  |  |  |  |  |  |  |
| Sex |  |  |  |  |  |  |  |  | 1.000 |
| Male | 8 | .. | .. | .. | 8 | .. | .. | .. |  |
| Female | 34 | .. | .. | .. | 34 | .. | .. | .. |  |
| Age | .. | 22 | 65 | 46.67 (11.38) | .. | 22 | 64 | 46.14 (11.69) | 0.809 |
| BMI | .. | 17.57 | 37.24 | 25.35 (4.58) | .. | 18.59 | 39.10 | 25.39 (5.34) | 0.758 |
| Medication** | 14 | .. | .. | .. | 0 | .. | .. | .. | 0.000 |
| PHQ-9 | .. | 1 | 25 | 10.73 (5.89) | .. | 0 | 10 | 3.71 (2.53) | 0.000 |
| SF-12 |  |  |  |  |  |  |  |  |  |
| Physical health | .. | 20.15 | 60.47 | 45.45 (10.60) | .. | 42.13 | 62.77 | 54.89 (4.18) | 0.000 |
| Mental health | .. | 21.08 | 60.76 | 40.58 (11.84) | .. | 24.93 | 62.12 | 52.16 (8.83) | 0.000 |
|  | Current MDD cases (*n* = 12) |  |  |  | Healthy controls (*n* = 12) |  |  |  |  |
| Current MDD |  |  |  |  |  |  |  |  |  |
| Sex |  |  |  |  |  |  |  |  | 1.000 |
| Male | 1 | .. | .. | .. | 1 | .. | .. | .. |  |
| Female | 11 | .. | .. | .. | 11 | .. | .. | .. |  |
| Age | .. | 26 | 61 | 49.92 (12.15) | .. | 26 | 61 | 49.92 (12.37) | 0.977 |
| BMI | .. | 17.63 | 31.80 | 24.43 (4.42) | .. | 18.87 | 39.10 | 26.47 (5.97) | 0.347 |
| Medication** | 7 | .. | .. | .. | 0 | .. | .. | .. | 0.000 |
| PHQ-9 | .. | 9 | 25 | 14.42 (5.40) | .. | 1 | 9 | 3.50 (2.50) | 0.000 |
| SF-12 |  |  |  |  |  |  |  |  |  |
| Physical health | .. | 21.63 | 57.26 | 39.13 (10.36) | .. | 43.55 | 61.83 | 53.98 (5.29) | 0.000 |
| Mental health | .. | 21.08 | 50.54 | 31.89 (9.48) | .. | 24.92 | 62.12 | 52.84 (10.77) | 0.000 |

Note: PDD: persistent depressive disorder, MDD: major depressive disorder, M: mean, SD: standard deviation, \**t*-tests, $\chi^2$ -tests and Mann-Whitney *U* tests were calculated depending on normality and scale of measurement, \*\* Medication: Psychopharmaceutical treatment.

While no significant difference in cell size (see supplementary Table 1) was detected for any disease group compared to healthy controls, cell deformability was altered in a disease-specific way. Two-tailed independent *t*-tests and Mann-Whitney *U* tests showed increased cell deformability in the granulo-monocyte cell fraction, especially in monocytes (*t*(58) = 3.105, *p* = 0.003) and neutrophils (*t*(58) = 2.887, *p* = 0.005) in participants with lifetime PDD compared to healthy controls (Figure 3). In current PDD an increased cell deformability in erythrocytes (*t*(28) = 2.082, *p* = 0.047), but no association with lymphocyte or granulo-monocyte deformability was detected (Figure 4). Current MDD (Figure 5) was associated with an increased cell deformability in lymphocytes (*U(22)* = 32, *p* = 0.0224), while we found no significant effect, but only trends towards increased cell deformability in lymphocytes and myeloid cells in lifetime MDD (Figure 6). For detailed statistics see Table 2.

**Table 2.**
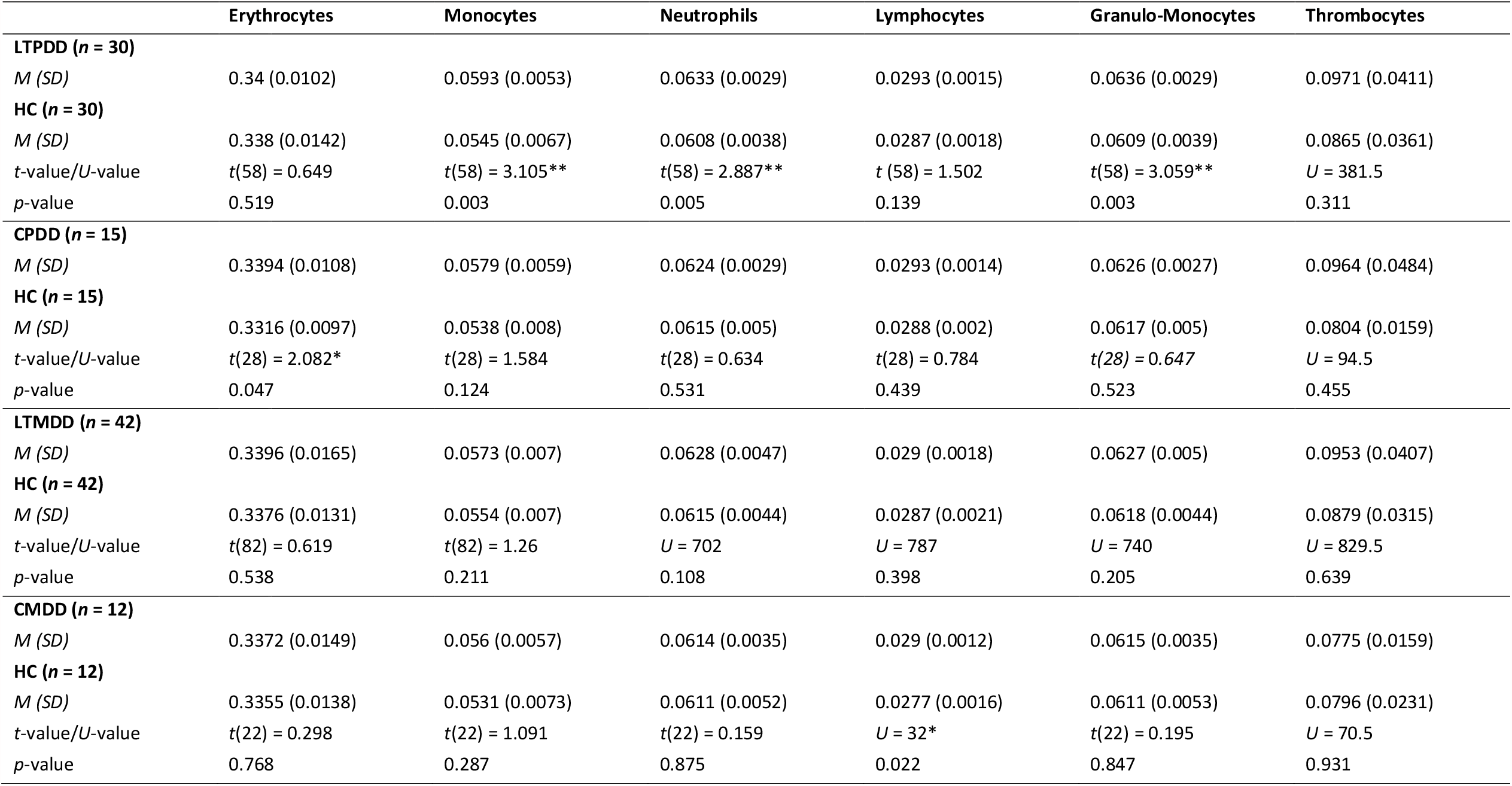
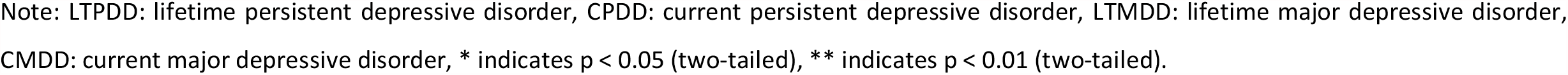
Mean cell deformability comparisons according to different diagnostic groups.

**Figure 3:**
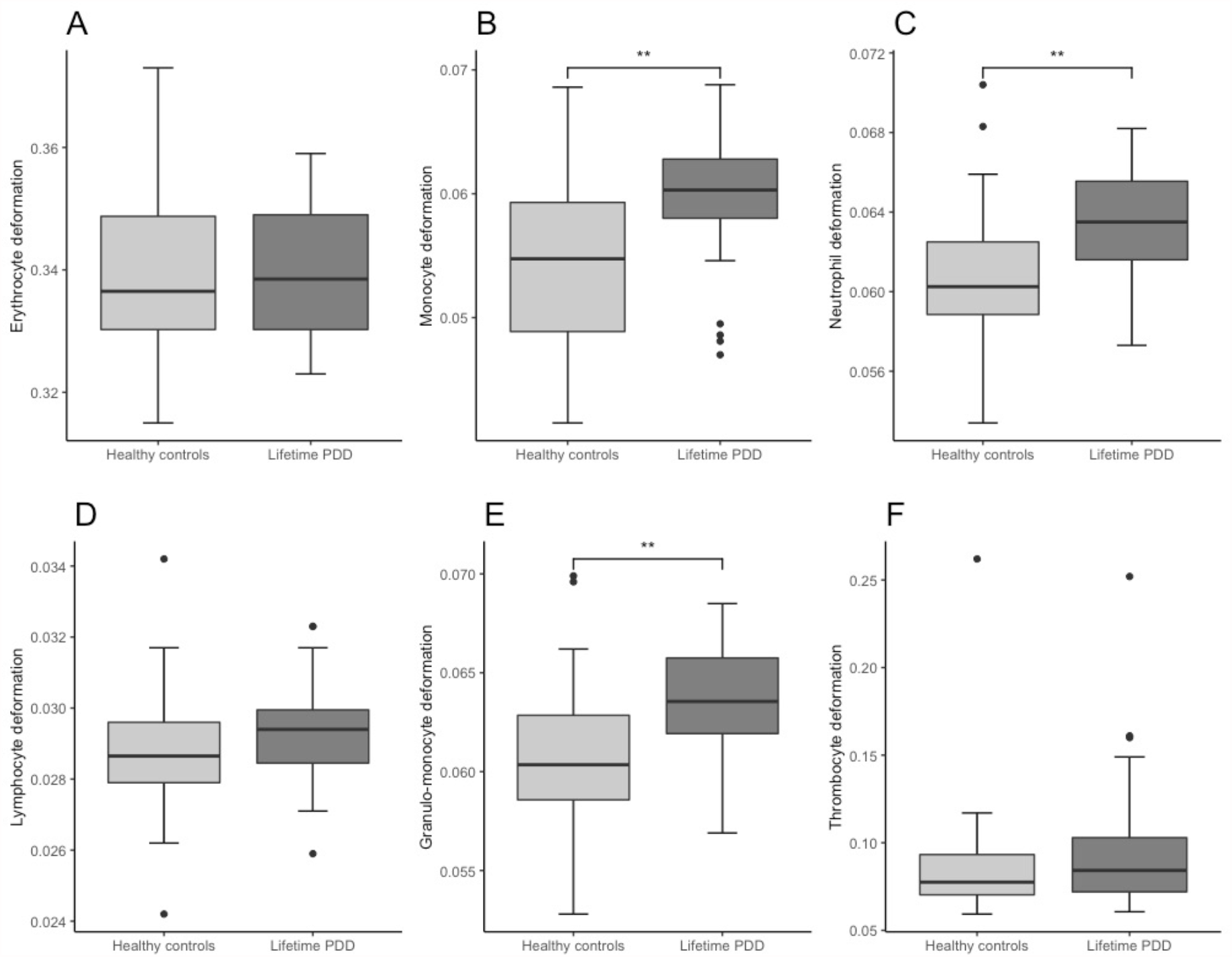
Combined boxplots for lifetime persistent depressive disorder (PDD) group and healthy controls regarding blood cell deformability. Box plots including interquartile range (boxes), median (lines), and range (whiskers). ** indicates significantly different deformability values at *p* < .01 (two-tailed).

**Figure 4:**
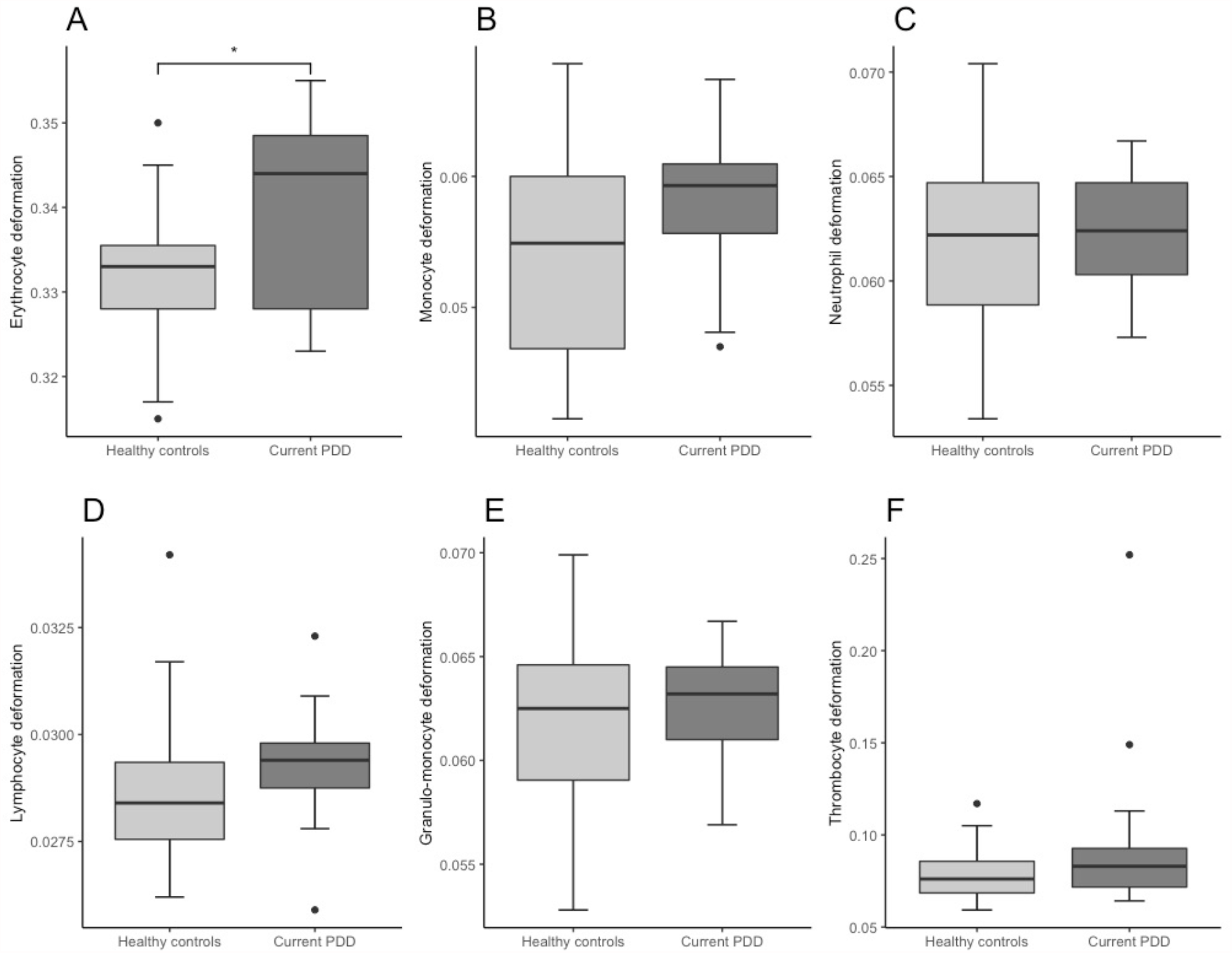
Combined boxplots for current persistent depressive disorder (PDD) group and healthy controls regarding blood cell deformability. Box plots including interquartile range (boxes), median (lines), and range (whiskers). * indicates significantly different deformability values at *p* < .05 (two-tailed).

**Figure 5:**
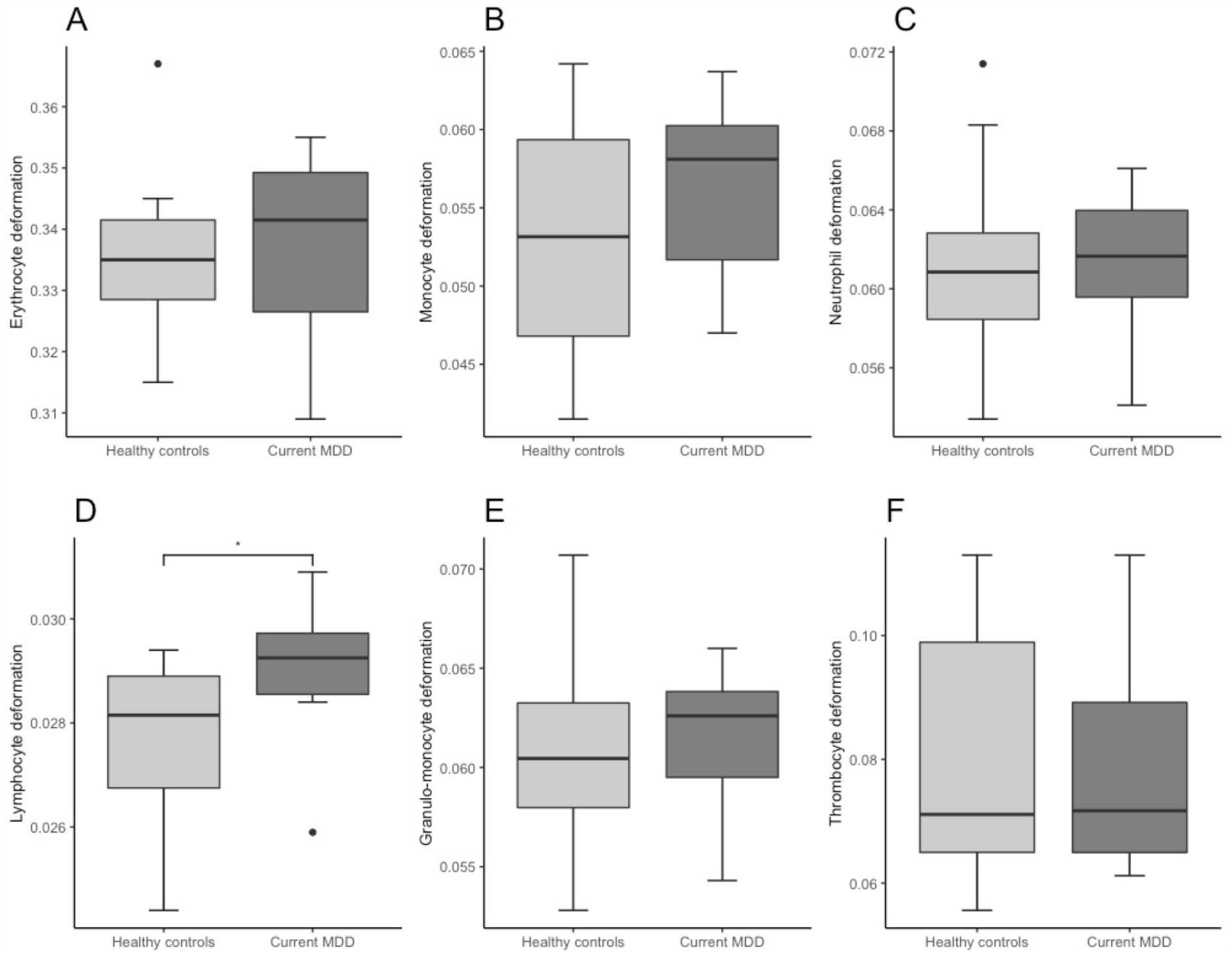
Combined boxplots for current major depressive disorder (MDD) group and healthy controls regarding blood cell deformability. Box plots including interquartile range (boxes), median (lines), and range (whiskers). * indicates significantly different deformability values at *p* < .05 (two-tailed).

**Figure 6:**
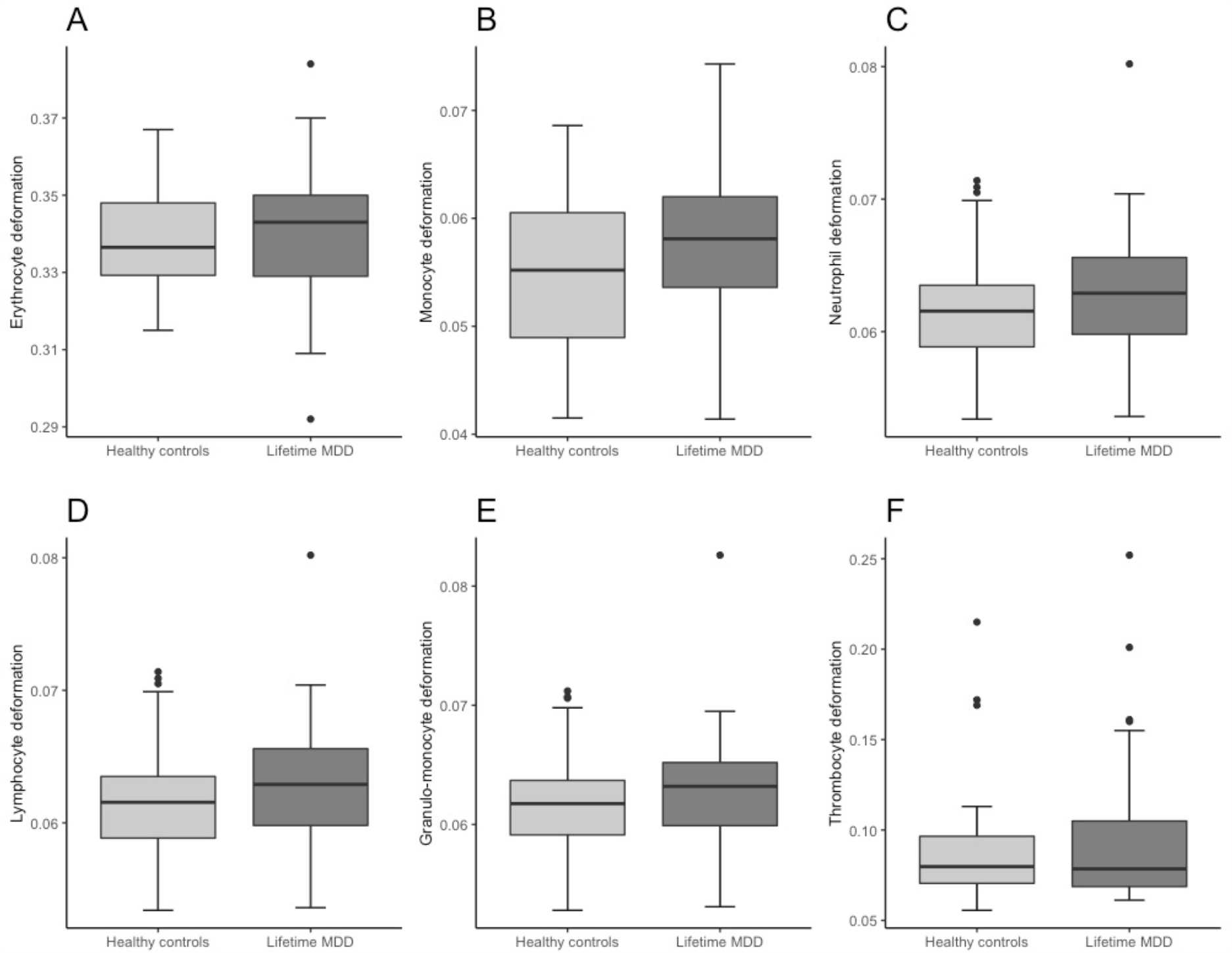
Combined boxplots for lifetime major depressive disorder (MDD) group and healthy controls regarding blood cell deformability. Box plots including interquartile range (boxes), median (lines), and range (whiskers).

Depressive disorder and healthy control groups showed no significant differences regarding age, sex, and BMI (see Table 1). Depressive disorder and healthy control groups significantly differed in PHQ-9 scores, SF-12 scores, and usage of psychopharmaceutic treatment (see Table 1). Psychopharmaceutical treatment was not associated with changes in cell deformability of erythrocytes (*t*(137) = -0.776, *p* = 0.439), monocytes (*t*(137) = -0.463, *p* = 0.644), neutrophils (*t*(137) = -0.603, *p* = 0.243), lymphocytes (*t*(137) = -0.096, *p* = 0.924), granulo-monocytes (*t*(137) = 1.138, *p* = 0.257), and thrombocytes (*t*(137) = -0.333, *p* = 0.3695). Subject questionnaire information on drug intake was used to create five groups according to medication: No medication (*n* = 75), psychopharmaceutical medication (*n* = 5), antihypertensive drugs (*n* = 16), thyroid dysfunction medication (*n* = 17), others (*n* = 4), and participants regularly taking in a combination of the preceding categories (*n* = 10). Univariate ANOVA showed no association between medication intake groups and cell deformability of erythrocytes (*F*_(5, 137)_ = 1.112, *p* = 0.352), monocytes (*F*_(5, 137)_ = 0.915, *p* = 0.474), neutrophils (*F*_(5, 137)_ = 0.774, *p* = 0.570), lymphocytes (*F*_(5, 137)_ = 1.051, *p* = 0.391), granulo-monocytes (*F*_(5, 137)_ = 0.904, *p* = 0.481), or thrombocytes (*F*_(5, 137)_ = 1.075, *p* = 0.377).

### Correlation analysis for diagnostic groups and matched controls

Table 3, 4, and 5 present Pearson, Spearman and partial correlations for the association between PHQ-9, SF-12 physical health, SF-12 mental health and cell deformability for each blood cell type. Partial correlations correcting for age, sex, BMI, and psychopharmaceutical treatment showed for the group consisting of lifetime PDD and matched control subjects significant correlations for higher depressive symptomatology and increased cell deformability for monocytes (*r*_*p*_ = 0.306; *p* = 0.022), neutrophils (*r*_*p*_ = 0.292; *p* = 0.028), and granulo-monocytes (*r*_*p*_ = 0.293; *p* = 0.028). In addition, current PDD and current MDD groups with matched controls showed significant positive partial correlations for depressive symptomatology and monocytes (*r*_*p*_ = 0.389; *p* = 0.050) and erythrocytes (*r*_*p*_ = 0.452; *p* = 0.046), respectively (see Table 3). No significant partial correlations emerged for the SF-12 physical health scale and cell deformability of any cell type (see Table 4). For the SF-12 mental health scale, as shown in Table 5, significant partial correlations with cell deformability emerged for the group consisting of lifetime PDD subjects and controls for neutrophils (*r*_*p*_ = -0.306; *p* = 0.022) and granulo-monocytes (*r*_*p*_ = -0.304; *p* = 0.022). Negative correlations indicate higher cell deformability being associated with lower self-reported mental health. No significant correlations with the SF-12 mental health scale were identified for cell deformability of the current PDD and matched control group as well as the lifetime MDD and matched control group. In the current MDD group and matched controls a significant partial correlation emerged for erythrocytes (*r*_*p*_ = -0.581; *p* = 0.008).

**Table 3.**
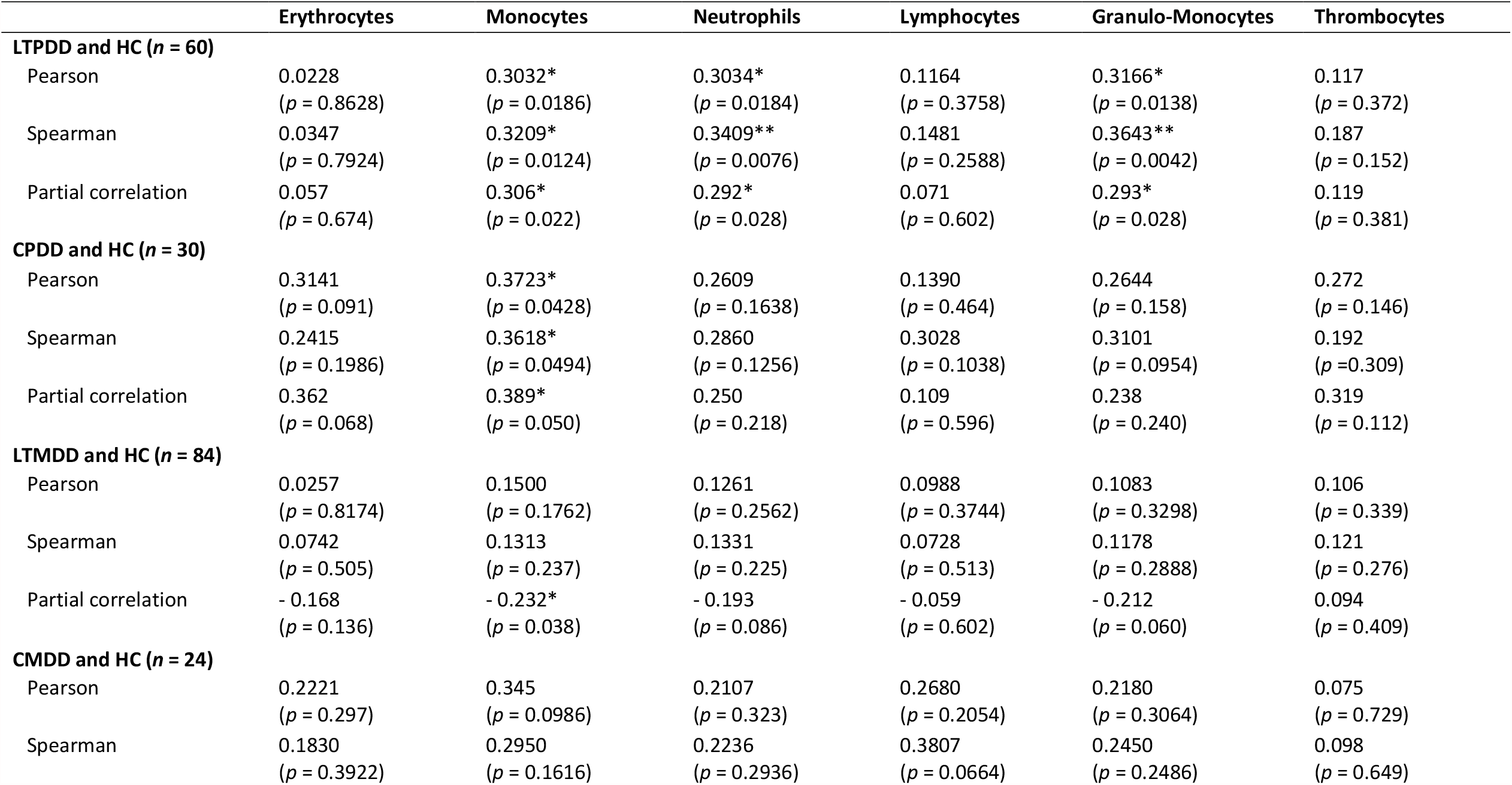
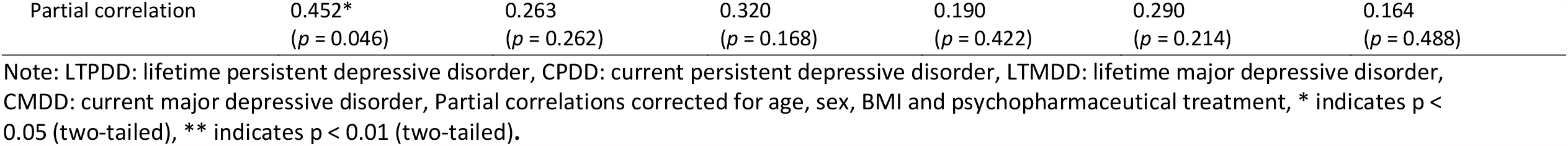
Association of depressive symptoms measured by PHQ-9 and cell deformability according to different diagnostic groups.

**Table 4.**
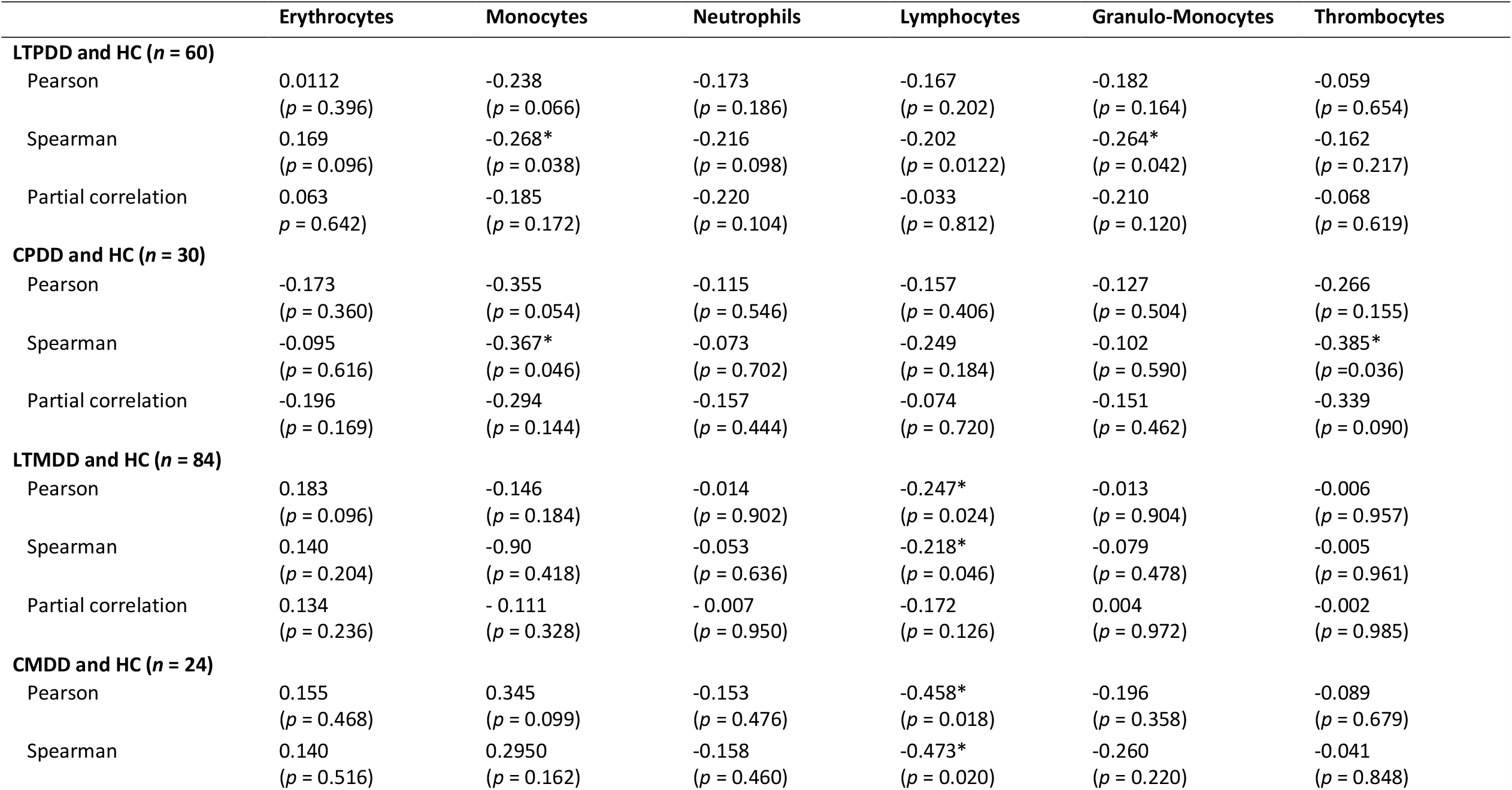
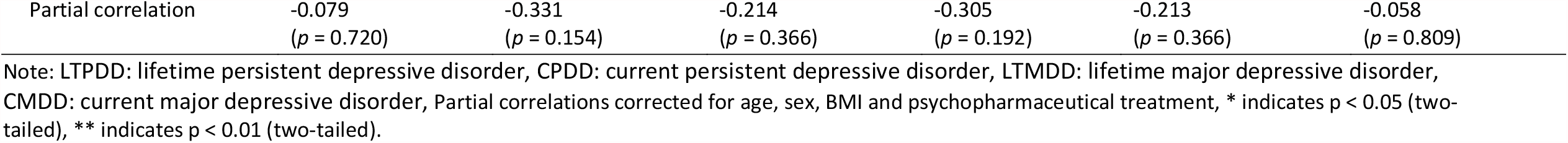
Association of physical health measured by SF-12 and cell deformability according to different diagnostic groups.

**Table 5.** Association of mental health measured by SF-12 and cell deformability according to different diagnostic groups.

|  | Erythrocytes | Monocytes | Neutrophils | Lymphocytes | Granulo-Monocytes | Thrombocytes |
| --- | --- | --- | --- | --- | --- | --- |
| **LTPDD and HC (n = 60)** |  |  |  |  |  |  |
| Pearson | -0.060<br>(p = 0.648) | -0.184<br>(p = 0.160) | -0.335**<br>(p = 0.008) | -0.132<br>(p = 0.314) | -0.342**<br>(p = 0.008) | -0.160<br>(p = 0.223) |
| Spearman | -0.071<br>(p = 0.588) | -0.162<br>(p = 0.216) | -0.342**<br>(p = 0.008) | -0.139<br>(p = 0.288) | -0.358**<br>(p = 0.002) | -0.241<br>(p = 0.063) |
| Partial correlation | -0.080<br>(p = 0.560) | -0.192<br>(p = 0.156) | -0.306*<br>(p = 0.022) | -0.135<br>(p = 0.320) | -0.304*<br>(p = 0.022) | -0.164<br>(p = 0.226) |
| **CPDD and HC (n = 30)** |  |  |  |  |  |  |
| Pearson | -0.292<br>(p = 0.116) | -0.283<br>(p = 0.130) | -0.355<br>(p = 0.054) | -0.212<br>(p = 0.262) | -0.335<br>(p = 0.054) | -0.276<br>(p = 0.140) |
| Spearman | -0.227<br>(p = 0.228) | -0.221<br>(p = 0.240) | -0.337<br>(p = 0.068) | -0.320<br>(p = 0.084) | -0.336<br>(p = 0.070) | -0.196<br>(p = 0.301) |
| Partial correlation | -0.314<br>(p = 0.118) | -0.295<br>(p = 0.144) | -0.330<br>(p = 0.100) | -0.225<br>(p = 0.268) | -0.321<br>(p = 0.110) | -0.284<br>(p = 0.160) |
| **LTMDD and HC (n = 84)** |  |  |  |  |  |  |
| Pearson | -0.111<br>(p = 0.314) | -0.121<br>(p = 0.274) | -0.146<br>(p = 0.186) | -0.081<br>(p = 0.466) | -0.127<br>(p = 0.252) | -0.169<br>(p = 0.127) |
| Spearman | -0.141<br>(p = 0.202) | -0.076<br>(p = 0.492) | -0.180<br>(p = 0.102) | -0.056<br>(p = 0.616) | -0.135<br>(p = 0.222) | -0.195<br>(p = 0.077) |
| Partial correlation | -0.066<br>(p = 0.558) | -0.131<br>(p = 0.248) | -0.118<br>(p = 0.298) | -0.090<br>(p = 0.426) | -0.107<br>(p = 0.344) | -0.153<br>(p = 0.179) |
| **CMDD and HC (n = 24)** |  |  |  |  |  |  |
| Pearson | -0.427*<br>(p = 0.038) | -0.191<br>(p = 0.370) | -0.255<br>(p = 0.230) | -0.150<br>(p = 0.486) | -0.250<br>(p = 0.238) | -0.241<br>(p = 0.256) |
| Spearman | -0.381<br>(p = 0.066) | -0.104<br>(p = 0.628) | -0.263<br>(p = 0.214) | -0.338<br>(p = 0.106) | -0.235<br>(p = 0.270) | -0.221<br>(p = 0.299) |
| Partial correlation | -0.581**<br>($p = 0.008$) | -0.127<br>($p = 0.592$) | -0.334<br>($p = 0.150$) | -0.185<br>($p = 0.434$) | -0.309<br>($p = 0.186$) | -0.405<br>($p = 0.077$) |

Note: LTPDD: lifetime persistent depressive disorder, CPDD: current persistent depressive disorder, LTMDD: lifetime major depressive disorder, CMDD: current major depressive disorder, Partial correlations corrected for age, sex, BMI and psychopharmaceutical treatment, \* indicates $p < 0.05$ (two-tailed), \*\* indicates $p < 0.01$ (two-tailed).

## Discussion

To our knowledge, this study is the first providing insights of the association between depressive disorders and cell morpho-rheological features of all major blood cell types. Our results suggest depressive disorders and in particular PDD to be associated with an overall increase in blood cell deformability, while for cell size no difference was observed. Hereby, the most consistent differences were found in lymphocytes, monocytes, and neutrophils highlighting the impact of depressive disorders on the mechanical properties of primary immune cells.

Morpho-rheological assessment of blood cells can provide crucial health status information, as changes in the cells mechanical constitution are associated with physiological or pathological function ^10^. Thus, increased blood cell deformability in individuals with depressive disorders compared to controls (see Fig. 3-6 and Table 3) provides novel insight into the pathophysiology of depressive disorders. However, due to the complex interplay between morpho-rheological features and cell function, we can only speculate on the underlaying causal link. A large body of evidence indicates depressive disorders to be associated with increased HPA-activity resulting in elevated cortisol levels ^26^. Moreover, a chronic low-grade inflammation with increased levels of proinflammatory cytokines and proteins such as interleukin-6 and C-reactive protein were described ^27^. Increased inflammatory signaling is known to interfere with the mechanics of myeloid cells ^19^. Only recently, these inherent biological underpinnings of depressive disorders have been linked to a dysregulation of the blood cell lipid composition leading to alterations in membrane structure ^6,28^. This altered membrane lipid composition is suggested to cause increased membrane bending and destabilization, potentially leading to an overall reduced integrity and altered functionality of blood cells ^6^. Thus, the association of increased cell deformability observed in depressive disorders is in accordance with current pathophysiological models of depressive disorders. In addition, Lynall et al. (2019) reported elevated numbers of immune cells in depressed individuals compared to controls. It is known that increased levels of glucocorticoids and catecholamines result in increased white blood cell count, as cells demarginate from the vessel walls. Interestingly, these observations were recently associated with cellular softening ^29^. In our study, elevated levels of circulating white blood cells in individuals suffering from depressive disorders cannot be confirmed, presumably due to the smaller sample size and the resulting lower power to detect the relatively small differences in immune cell count. However, increased immune cell deformability in depressed patients might be a direct response to cortisol levels and can be indicative for immune cell activation, as suggested by chronic low-grade inflammation. On the other hand, we also found increased erythrocyte deformability in individuals with current PDD. A tight control of homeostatic erythrocyte deformability is arguably of high importance in order to provide passage through narrow capillaries and tissue oxygen supply, critical for various organs including the central nervous system. Whether an altered lipid composition interferes with erythrocyte oxygen transport due to increased erythrocyte deformability or whether increased erythrocyte deformability is a body response to keep oxygen supply constant needs to be further examined. Our results highlight altered blood cell morpho-rheological properties in depressed patients to play a critical role in the pathophysiological processes; however, to what extend blood cell deformability is involved in the disease progression is yet far from understood. Furthermore, we did not identify differences in cell size of any of the investigated blood cells underlining the potential to readout cell deformability as indicator for altered cell function. Importantly, since participants were not drug naïve, the potential association of specific medication types such as psychopharmacological agents, thyroid dysfunction medication, or antihypertensives with cell deformability was examined. No association was identified for cell deformability with any specific medication type suggesting that depressive disorders and not the examined medication types affect cell deformability.

Another point to be considered is that there are different ways to assess cell deformability. While the applied RT-DC method represents a versatile tool to measure specifically blood cell deformability on short time scales ^10,18^, other approaches provide important insights into cell mechanical properties on longer time scales. A recent study reports a reduced erythrocyte deformability in 16 patients with myalgic encephalomyelitis / chronic fatigue syndrome compared to age-matched healthy controls ^15^. They examined how long erythrocytes take to cross a 5 μm x 5 μm channel at a negative pressure of -13.79 kPa. The cells deform tactile to fit through the channel, which took approximately 13 ms. This is more than three times as long as cells are deformed in RT-DC. Thus, in the study by Saha et al. (2019), the cell viscous properties might play a more pronounced role in order to pass through the channel compared to RT-DC. Interestingly, the erythrocytes derived from myalgic encephalomyelitis / chronic fatigue syndrome patients were found to be larger compared to healthy controls. Indicating again a disease-specific alteration of the morpho-rheological features. A similar report applying centrifugation of erythrocytes through a filter of 5 µm pores quantifies erythrocyte deformability as ratio of filtered erythrocytes to initial cell number. They examined 54 children with autism spectrum disorders and identified impaired erythrocyte deformability associated with more severe restricted and repetitive symptomatology ^16^.

## Strengths and Limitations

Strengths of our work are the pioneering character of the blood cell deformability measurement carried out using RT-DC in individuals with depressive disorders and age- and sex-matched healthy controls and the acquisition of an average of 115,000 cell images of each of the 139 subjects representing a yet not achieved large dataset. For fast, automated cell classification of the resulting dataset of over 16 million images, an artificial intelligence-based analysis was leveraged (Figure 1). It needs to be noted, that blood samples were immediately measured using RT-DC within a time frame of three hours after sampling since longer storage times or freezing the cells could potentially alter cell membrane and deformability properties. In addition, the standardized clinical evaluation of the individuals provides the highest level of diagnostic validity and reliability. A limitation of our study is the fact that the participants were not drug naïve. However, since we intended to investigate depressive disorders in general including individuals with a lifetime or current PDD or MDD diagnosis, it is very difficult to include drug naïve individuals only. Therefore, in our analyses, we consistently controlled for the potential influence of medication on cell deformability. Another limitation is that PDD and MDD groups partially overlap raising the question whether findings are related to the combination of disorders, to the general severity of the symptoms of depression, or to chronicity. Further, the relatively low number of male individuals in our study renders a generalization to the male population difficult.

## Conclusions

To conclude, this study provides to our knowledge the first evidence of a relationship between peripheral blood cell deformability and depressive disorders. As the pathophysiology of depressive disorders is only poorly understood, and HPA-hyperactivity and chronic low-grade inflammation represent landmarks of the current pathophysiological model, our results further point toward a persistent activated immunity in depressive disorders. In combination with altered lipid metabolism and blood cell membrane assembly, cell functional changes mediated by cytoskeletal adaptations are very likely to occur. In agreement with other reports, we found, that these cell functional changes can be detected disease-specific by morpho-rheological measurements, potentially leading to a co-diagnostic marker. Thus, our study broadens the understanding of the current physiological underlying causes of depressive disorders in blood cells and delivers a clinical grade method to assess erythrocyte and immune cell functionality. Future research will be needed to confirm our findings in larger cohorts, in order to render the discriminant potential of cell morpho-rheological properties in depressive disorders more specific. Moreover, the potential to reverse increased peripheral blood cell deformability might be harnessed to develop new pharmacological treatments restoring optimal levels of cell deformability, and cell function and thereby reduce depressive burden.

## Data Availability

All data will be made available upon request. Data will be deposited on OSF after study completion and publication of manuscripts.

## Contributors

AW and MK designed the study, performed experiments and data analysis, and co-wrote the manuscript. AMK, JE, and MH performed experiments and data analysis and co-wrote the manuscript. CH and LDW supported data analysis. CK, JG, and KBB provided funding, infrastructure/equipment, and co-wrote the manuscript.

## Declaration of interest

Christoph Herold owns shares of, and is employed at Zellmechanik Dresden GmbH, a company selling devices based on real-time deformability cytometry. Zellmechanik Dresden GmbH did not have any role in the conception and planning of this study, or its preparation for publication. All other authors state that they have no actual or potential conflict of interest to declare, including any financial, personal, or other relationships with other people or organisations within 3 years of beginning the submitted work that could influence or bias their work.

## Role of the funding source

- Research pool TU Dresden (F-004242-552-848-1040103) awarded to AW
- Faculty of Psychology of the TU Dresden (MK201911) awarded to AW
- Alexander von Humboldt Professorship awarded to JG
- German Research Foundation (DFG) – (399422891) awarded to MH and JG
- Federal Ministry of Education and Research (BMBF) (01ER1307) awarded to KBB

The funding sources are national and university funding sources and had no influence on the writing of the manuscript or the decision to submit it for publication. None of the authors received financial incentives from industrial companies to write this publication. The corresponding author had full access to all study data and had the final responsibility for the decision to submit the paper for publication.

## Notes

### Clinical Trial

No registration was performed since the present study is not an interventional but an observational study (case-control comparison).

### Author Declarations

All subjects signed consent forms regarding data privacy, clinical data collection and saving, and blood extraction. The study was approved by the local ethics committee of the Dresden University of Technology.

