## Supplementary for "Depressive disorders are associated with increased peripheral blood cell deformability: A cross-sectional case-control study (Mood-Morph)"

### Supplementary Material

**Supplementary Table 1. Mean cell area size comparisons according to different diagnostic groups**

| | Erythrocytes | Monocytes | Neutrophils | Lymphocytes | Granulo-Monocytes | Thrombocytes |
|---|---|---|---|---|---|---|
| **LTPDD (n = 30)** | | | | | | |
| M | 40.9390 | 66.1913 | 67.7062 | 37.6273 | 67.7894 | 5.6540 |
| SD | 1.2461 | 2.3062 | 1.1810 | 1.2022 | 1.1940 | 0.8728 |
| **HC (n = 30)** | | | | | | |
| M | 40.8469 | 66.0337 | 67.6605 | 37.9806 | 67.7738 | 5.5787 |
| SD | 1.9207 | 2.6292 | 1.2462 | 1.7160 | 1.2085 | 6.8770 |
| *t*-value | - 0.220 | - 0.247 | - 0.146 | 0.924 | - 0.050 | - 0.333 |
| *p*-value | 0.4135 | 0.403 | 0.4425 | 0.180 | 0.480 | 0.3700 |
| **CPDD (n = 15)** | | | | | | |
| M | 40.7275 | 65.9237 | 67.7512 | 37.5837 | 67.9206 | 5.6507 |
| SD | 1.1010 | 2.5406 | 1.2405 | 1.2379 | 1.1676 | 1.1016 |
| **HC (n = 15)** | | | | | | |
| M | 40.7029 | 66.4452 | 67.9298 | 37.8058 | 68.0457 | 5.4427 |
| SD | 1.2413 | 3.0257 | 1.4592 | 2.1836 | 1.4232 | 0.5773 |
| *t*-value | - 0.057 | 0.511 | 0.361 | 0.343 | 0.263 | - 0.648 |
| *p*-value | 0.4775 | 0.3065 | 0.3605 | 0.367 | 0.397 | 0.2615 |
| **LTMDD (n = 42)** | | | | | | |
| M | 41.0876 | 66.4817 | 68.0362 | 37.8409 | 68.0979 | 5.4017 |
| SD | 1.4583 | 2.2434 | 1.5556 | 1.1895 | 1.4886 | 0.6913 |
| **HC (n = 42)** | | | | | | |
| M | 40.7902 | 66.2198 | 67.9257 | 37.7184 | 68.1101 | 5.5390 |
| SD | 1.5475 | 2.3655 | 1.5385 | 1.5029 | 1.4816 | 0.9077 |
| *t*-value | - 0.906 | - 0.521 | - 0.327 | - 0.414 | 0.038 | 0.780 |
| *p*-value | 0.1835 | 0.302 | 0.372 | 0.34 | 0.485 | 0.2185 |
| **CMDD (n = 12)** | | | | | | |
| M | 40.7546 | 65.8923 | 67.5383 | 37.5598 | 67.6076 | 5.1817 |
| SD | 1.4692 | 1.9439 | 1.1775 | 1.3376 | 1.0862 | 0.8033 |
| **HC (n = 12)** | | | | | | |
| M | 41.0904 | 65.8106 | 67.6602 | 37.9470 | 67.7980 | 5.5175 |
| SD | 1.4585 | 2.6542 | 1.7148 | 1.2257 | 1.7581 | 0.6739 |
| *t*-value | 0.562 | - 0.086 | 0.203 | 0.739 | 0.319 | 1.109 |
| *p*-value | 0.29 | 0.466 | 0.4205 | 0.234 | 0.3765 | 0.1395 |

Note: LTPDD: lifetime persistent depressive disorder, CPDD: current persistent depressive disorder, LTMDD: lifetime major depressive disorder, CMDD: current major depressive disorder, Mean area size ($\mu m^2$), * indicates $p < 0.05$ (one-tailed)
